# Convergence of Comorbidity and COVID-19 Infection to Fatality: An Investigation Based on Health Assessment and Vaccination among Older Adults in Kerala

**DOI:** 10.1101/2021.01.06.20249030

**Authors:** Sindhu Joseph, Jijo Pulickiyil Ulahannan, A J Parvathy

**Affiliations:** Department of Travel and Tourism Management, Govinda Pai Memorial Government College, Kannur University, Kerala, India; Department of Physics, Government College Kasaragod, Kannur University, Kerala, India; Department of Computer Science, Vellore Institute of Technology, Vandalur - Kelambakkam Road, Chennai, Tamil Nadu, India, 600127; Collective for Open Data Distribution-Keralam (CODD-K)

**Keywords:** Comorbidity, Convergence, COVID-19 Mortality, Fatality, Older population, Vaccination

## Abstract

**Objective:** To investigate the impact of age, comorbidity, and vaccination in the fatality of older COVID-19 patients in the state of Kerala, India, based on their comorbidity and vaccination status.

**Methods:** It is a cross sectional study adopting a mixed method approach conducted among the older population in Kerala. To study the health profile, 405 older people were surveyed, and 102 people were interviewed in-depth at their households, between June to November 2020. The results of the study were triangulated with elderly COVID-19 fatality data, available from the citizen-science dashboards of the research team and Department of Health, Kerala. Vaccination data was retrieved from cowin.gov.in to study its impact. The data was analysed using the IBM SPSS version 22.0.

**Results:** Age is a predictor of COVID-19 fatality. Diabetes, hypertension, heart diseases, CKD and COPD are the significant predictors of elderly COVID-19 fatality. The current comorbidity profile of the total older population matches with the comorbidities of the COVID-19 elderly death cases. Vaccination has impacted COVID-19 mortality after vaccinating 65 percent (first dose) of the elderly.

**Conclusions:** Age and comorbidities can predict potential fatality among older COVID-19 patients. Timely and accurate health data and better knowledge of high-risk factors such as comorbidity can easily guide the healthcare system and authorities to efficient prevention and treatment methodologies. Knowledge on prevailing NCDs can drive early preparedness before it converges with an epidemic like the present zoonotic disease. Priority must be given for elderly vaccination to bring down the mortality rates.

## INTRODUCTION

It is already acknowledged that health conditions deteriorate when age increases and ageing goes hand in hand with many behavioural issues and disabilities, contagious infections, lifestyle, and chronic diseases. Therefore, the impact of multi-morbidity on various aspects such as quality of life, functionality and risk of mortality becomes a matter of present discussions worldwide. They face health issues such as loss of appetite, gastric and body pain, cataract, arthritis, uterine problems, hypertension, diabetes mellitus, general weakness, stroke, muscular-skeletal and respiratory diseases [1–3].

In India, geriatric care has been adversely affected by the population size leading to inadequacy of the public healthcare system in providing equitable healthcare. They face NCDs’ threat, including cancers, CVD (cardiovascular disease), respiratory diseases, and diabetes and one out of every two older people suffers from at least one chronic disease requiring life-long medication, particularly in urban areas [4].

Kerala has increased one million older population every consecutive year since 1981 [5]. If this trend continues incessantly, it is expected to surpass the proportion of young and old in between 2021 and 2031 [5]. In 2020, 48 lakh people of Kerala were above 60 years of age of which 15 percent of them were above 80 years, the fastest-growing group among the old. By 2025, about 20% of the population in Kerala [6].

Kerala has the highest outpatient doctor consultation per 1,000 population compared to the rest of the states in India [7]. Higher hospitalisation rates indicate higher morbidity levels [8]. In a 2004 study, about 25% of the older people reported having poor health status [9]. Their common ailments were paralysis, urinary problems, CVD and cancer in 2010 [10], hypertension, diabetes mellitus, cataract and heart disease in 2011 [8], CVD, diabetes, musculoskeletal, and respiratory disease in 2016 [11] and diabetes, abdominal obesity, and hypertension in 2016-17 [12]. In 2020, about 70 lakh older people will have cancer, diabetes, hypertension, respiratory tract infections, kidney, and heart diseases [13]. Based on these previous studies, it can be assumed that the significant prevalent diseases of older people in Kerala are hypertension, diabetes, CVD, cancer, and respiratory diseases.

## THE COVID-19 FATALITY

Kerala reported the first COVID-19 case in India in January 2020 and was the first state that saw the first wave of the disease spread in the country. The spread of COVID-19 affected mostly the older people. Patients with chronic comorbidities, including malignancy, CVD, diabetes, hypertension, kidney and respiratory disease are prone to the fatal outcome of COVID-19 infection [14–21]. Various studies [14–16] found that for CVD and hypertension, the use of renin-angiotensin system inhibitors may accelerate the susceptibility to SARS-CoV-2 infection.

Effective and safe vaccines are the pharmaceutical interventions to prevent this pandemic and different types of vaccines are getting acceptance in different regions. The vaccination for the Age-Appropriate category (persons over 60 years of age, and persons between 45 and 59 years with comorbid conditions) started from March 1, 2021, onwards. The public over 45years of age started on April 1, 2021, and 18-44 group vaccination started on May 1, 2021. Hence, we considered mortality data after April 1 for comparison with that of pre vaccination data.

Despite the outnumbered post-COVID-19 studies describing comorbidities and the poor clinical outcomes leading to fatality, the results are inconsistent. Throughout the COVID-19 outbreak, wide variations in CFR (Case Fatality Rate) and IFR (Infection Fatality Rate) estimates have been noted, which are misleading [17]. There is a dearth of studies on COVID-19 and comorbidity while having timely data on the health profile of a population. Likewise, the study findings related to the health status, comorbidity profile, and the NCDs scenario of the older people over a decade in Kerala are also inconsistent, [8,10–13] and there is no study on the impact of vaccination among the older population in Kerala. In this context, this study is intended to fill the above gaps and plan to relate the comorbidity profile of the older people in Kerala in the post-COVID-19 scenario to the elderly mortality rate of COVID-19, which will help to identify the severity of risks. It is essential to draw a clear picture of the health status and morbidity levels of the older people amidst the pandemic to assess the possible implications of comorbidities while it converges with infectious diseases.

This study is unique as it measured the comorbidity level of the older people, while the pandemic is ongoing with simultaneous assessment of the older people COVID-19 deaths due to the suspected comorbidity levels. Even though it is too early to assess the impact of vaccination, the study attempted to show its impact on COVID-19 elderly mortality. To check and identify a consistent pattern in the association between age and COVID-19 mortality, an extensive review of the literature was carried out. The study insights will help to frame geriatric treatment policies and preventive measures while NCDs converge with infectious diseases.

## METHODOLOGY

This study is based on a mixed methodology approach. The study used a concurrent timing strategy where both the quantitative and qualitative strands occurred during a single phase, from June 2020 to November 2020. Quantitative data was collected from 405 older people from 36 panchayats of Kerala, adopting multi-stage sampling techniques, using a structured questionnaire. The study area was divided into three zones, South, Central and North Kerala. From each zone, three districts were selected at random. Four panchayats from each district were selected by drawing lots, making 36 panchayats of Kerala under study. From each selected panchayath, 11 households with older people were selected randomly.

The survey questionnaire included questions on socio-demographic profile and the disease profile of the elderly. The respondents were asked to mark appropriate responses against the names of different diseases in a 5-point scale ranging from ‘Always to Never’. The data were analysed with the IBM SPSS version 22.0. The percentage of a particular disease is calculated exempting the ‘never’ and ‘rarely’ responses, therefore including those who suffer the disease ‘always’, ‘frequently’ and ‘sometimes.’

Using a panchayat-wise list received from the *Vayomithram* coordinators (a government project for holistic care of older people), Anganvadi and *Asha workers* (health workers), older people above 60 years of age were selected randomly for the qualitative interview. The interviews were conducted at their households for about 30 - 45 minutes. An average of seven interviews per district were conducted and therefore 102 in-depth interviews were undertaken from all the fourteen districts of Kerala. They were asked about the present ailments and health status. To further enrich the data, 12 interviews were conducted with *Vayomithram (a project for elderly healthcare in the public sector)* district coordinators of 12 districts in Kerala. Analysis of data used a grounded theory research design, where the approach intends to develop concepts, insights, or models grounded in the participant’s data [18].

Directorate Health Services (DHS) website (http://dhs.kerala.gov.in) gives updates about the COVID-19 spread. However, access to primary data is not complete, and therefore, a multi-disciplinary team of experts compiled data through a citizen science initiative, managed by the researchers of this study (https://doi.org/10.5281/zenodo.3818096) [19,20]. A systematic analysis of the above mentioned two dashboards on CFR and comorbidities was done. The data was extracted from the death cases reported in Kerala with COVID-19 infection between January 2020 and May 19, 2021 (Fig. 1). Vaccination data was retrieved from the centralised portal of India (cowin.gov.in). The quantitative and qualitative strands were triangulated and integrated at the point of interpretation and drawn inferences from them.

**Figure 1.**
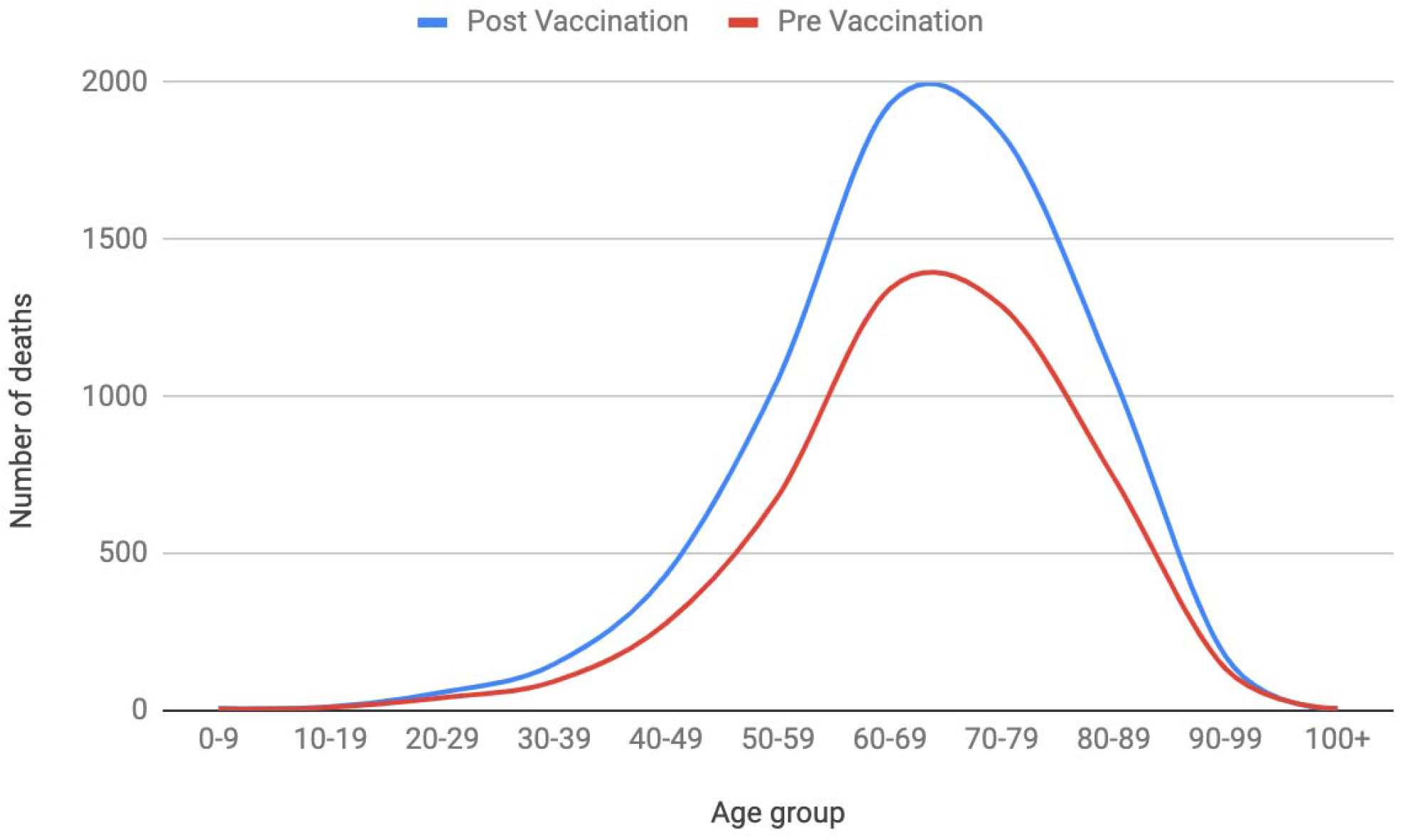
Covid-19 Deaths in Kerala-Age wise till May 19, 2021 Source: DHS, Government of Kerala, COVID-19 Dashboard

The study is conducted as per the guidelines of ICSSR (Indian Council of Social Sciences) New Delhi, India under the ICSSR – IMPRESS scheme (F. No. IMPRESS/P1132/428/2018-19/ICSSR). The sanctioned study on geriatric health was carried out while the pandemic is ongoing, enabling the research team to simultaneously investigate older people COVID-19 deaths due to the suspected comorbidity levels. This study adopted the definition of WHO [21] for COVID-19 death. Before the qualitative and quantitative data collection, the participants were briefed about the study’s purpose, and informed consent was obtained. COVID-19 protocol was adhered to while collecting the data.

## RESULTS

The socio-demographic profile (Table 1 supplementary data) shows that 55.3% were males, and 44.7% were females. Around 50.1% were in the 60-69 age category, followed by 31.6% in the 70-79 category.

**Table 1.**
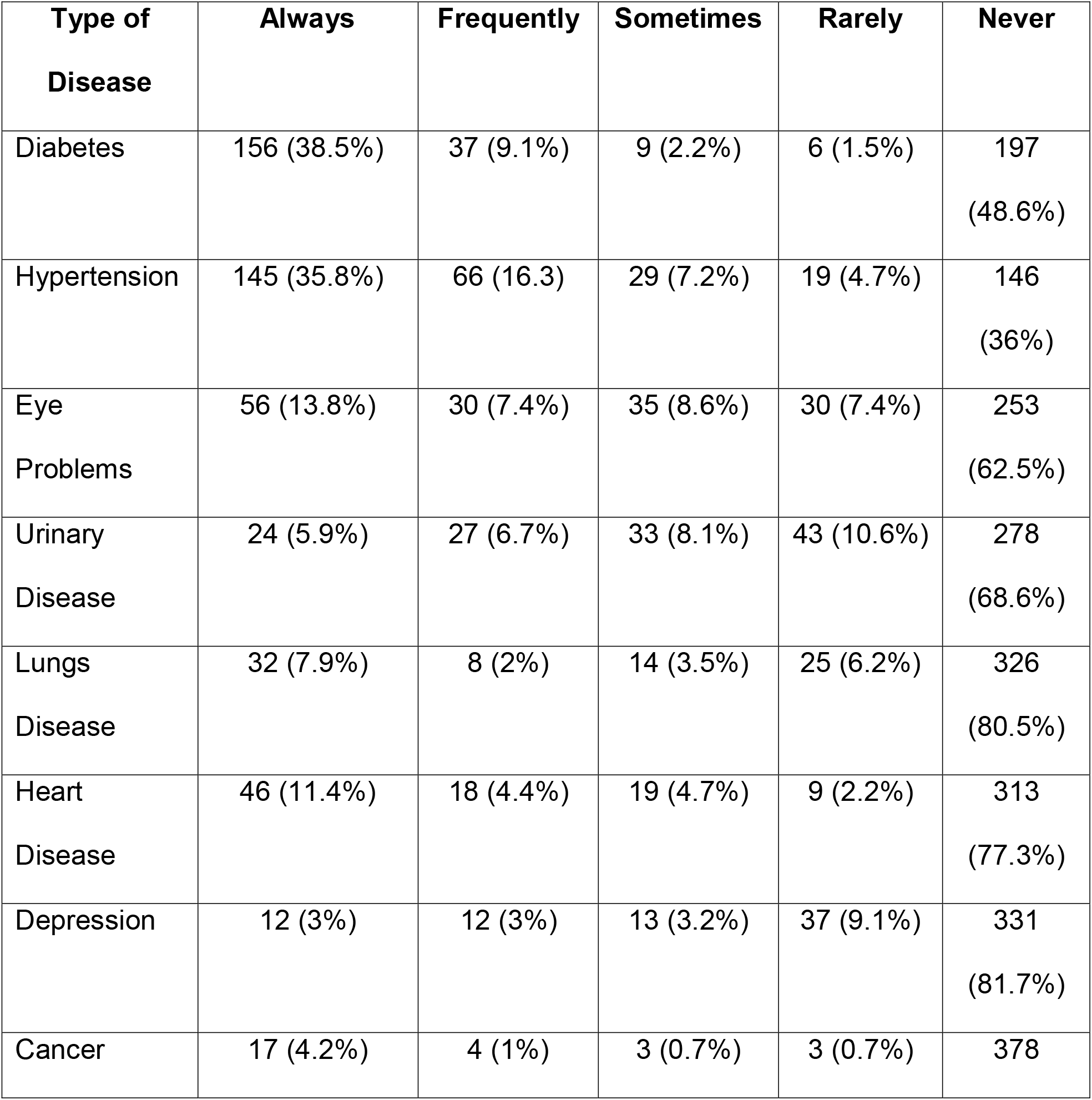
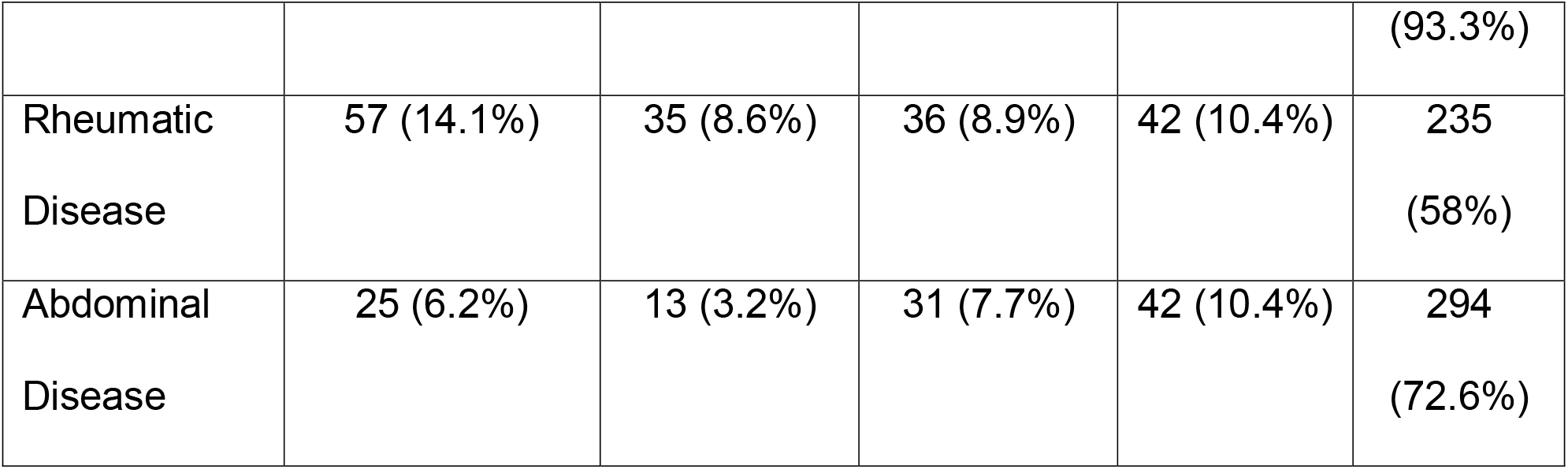
Elderly Disease Profile in Kerala.

## COMORBIDITY OF THE ELDERLY IN KERALA

Table 1 shows that older people of Kerala suffer from Hypertension (59.3%), Diabetes (49.8%), heart disease (20.5%), lungs disease (13.4%), Cancer (5.9%), Rheumatic Disease (31.6%), Eye Problems (29.8%), Urinary Disease (20.7%), Abdominal Disease (17.1%) and Depression (9.2%). Many of them have two or more diseases (comorbidities). Interviews also showed that most of the older people had lifestyle diseases like Hypertension, Diabetes, Heart Disease, Cancer, and Cholesterol (Table 2 supplementary data). *Vayomithram* coordinators also shared the same view.

**Table 2.** Comorbidity of the COVID-19 death cases.

| Comorbidity | AUG | Sept | Oct | Nov | Dec | Jan | Feb | Total |
| --- | --- | --- | --- | --- | --- | --- | --- | --- |
| Diabetes mellitus | 120 | 130 | 173 | 287 | 565 | 708 | 317 | 2300 (55.89%) |
| Hypertension | 116 | 119 | 169 | 302 | 536 | 661 | 313 | 2216 (53.85%) |
| CAD | 54 | 57 | 89 | 130 | 252 | 308 | 130 | 1020 (24.78%) |
| CKD | 36 | 52 | 103 | 96 | 178 | 221 | 96 | 782 (19%) |
| CVA | 17 | 22 | 20 | 33 | 80 | 101 | 36 | 309 (7.50%) |
| COPD | 23 | 22 | 34 | 63 | 98 | 130 | 71 | 441 (10.71%) |
| Cancer | 15 | 16 | 19 | 27 | 53 | 45 | 26 | 201 (4.88%) |
| Bedridden | 5 | 16 | 3 | 5 | 3 | 11 | 6 | 49 (1.19%) |
| CLD | 8 | 12 | 6 | 14 | 29 | 44 | 19 | 132 (3.20%) |
| Bronchial asthma | --- | 3 | 4 | 16 | 11 | 22 | 6 | 62 (1.50%) |
| TB | --- | 1 | 6 | 14 | 16 | 18 | 9 | 64 (1.55%) |
| No comorbidities | 2 | 2 | 12 | 18 | 51 | 69 | 30 | 184 (4.47%) |
| Total deaths | **223** | **230** | **302** | **519** | **998** | **1258** | **585** | **4115** |

### Covid-19 Deaths in Kerala-Age wise

Fig. 1 shows that age is a predictor of increased mortality rate. Till May 19, 2021, 6724 COVID-19 deaths happened in Kerala, of which 5012 (74.54%) deaths were older people, where the majority (28.72%) belongs to the 60-69 age group (Fig.1).

### Elderly COVID-19 Deaths and Comorbidity

Since the comorbidity data is not available for the elderly alone, the study used the comorbidity data of the total death cases of COVID-19. Death Audit report is available till August 28, 2021, published by the Department of Health, Kerala. As of February 28, 2021, out of the 4115 people who died, 3931 (95.53%) of them had comorbidities (Table 2 and Fig. 2). Major comorbidities are diabetes (55.89%), hypertension (53.85%), CAD (24.78%), CKD (19%), COPD (10.71%) and only 4.47% had no comorbidities (Table 2 and Fig 2).

**Figure 2.**
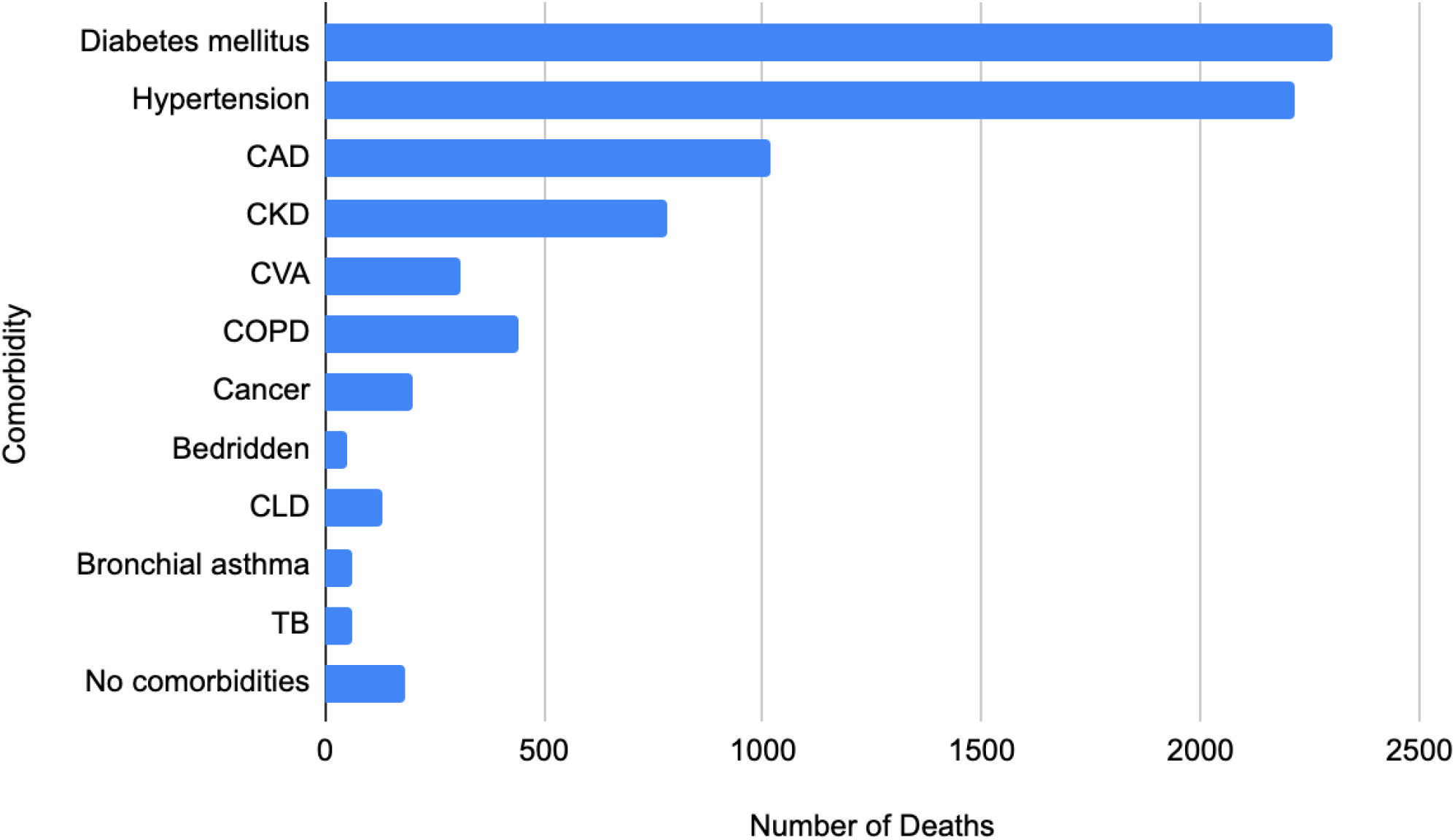
Types of Comorbidity Prevalent in Fatality Cases of COVID-19. Source: Death Audit Reports up to February 2021, DHS, Government of Kerala (Department of Health & Family Welfare, March 18, 2021)

The effect of this pandemic can be studied by using CFR, CMR (Crude Mortality Rate), and IFR [17,22]. CFR helps to recognise the disease severity, risks, and healthcare system quality [17]. CFR and recovery rates are important indicators during epidemics and pandemics which will help clinicians in stratifying patients in terms of the extent of care required, and in turn, increase the possibilities of survival from the deadly pandemic [23]. The CFR of an ongoing pandemic is calculated using the formula [17].

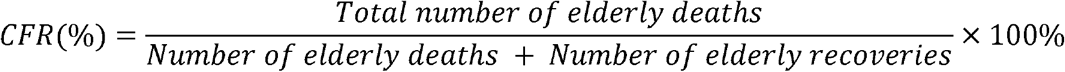

As per this formula, the CFR of the older people COVID-19 death cannot be estimated as the number of infected and recovered older people data is not available. Hence, as per the available published COVID-19 epidemiology data on total number of infections and recoveries, in Kerala, the CFR=0.35%, IFR=0.30%, CMR = 0.02% till May 19, 2021 (Table 3). Here CMR estimates the probability of any individual in a population dying from the disease [17,22] and IFR estimations give the proportion of fatality among all infected.

**Table 3.** COVID-19 Indicators (CFR, CMR, IFR) Note: The 2021 Projected population of Kerala is 3,54,89,000 as per the report of National Commission on Population of India. The number of the elderly population is projected as 52,71,660.

| **Type of data** | **No. of infected cases** | **No of elderly with comorbidity** | **No of deaths** | **CFR %** | **CMR (x1000 00)** | **IFR %** |
| --- | --- | --- | --- | --- | --- | --- |
| DHS Dashboard (as on 19 May 2021) | 2233468 | Data is available till Feb 28, 2021 | 6724 | 0.35 | 18.94 | 0.30 |
| DHS Dashboard (as on 28 February 2021) | 1059403 | 4115 | 4197 | 0.49 | 11.83 | 0.40 |

### Impact of Vaccination

Total vaccinations given in Kerala is 65,71,970 (18.52%) as on May 19, 2021, of which the number of vaccinated elderly (first dose only) is 33,08,148 (62.75%). Though the vaccination was started only from March 1, 2020, slight decline in elderly mortality has been noticed between the pre-vaccination and post-vaccination period (Fig. 1).

## DISCUSSION

The emergence of the COVID-19 disease has been spread with surges and resurges. In this context, this paper is intended to associate older people’s health scenario with COVID-19 comorbidity and fatality rates, using a concurrent mixed-method approach.

Comorbidity profile of older people, age-wise death report, and the type of comorbidities of COVID-19 death cases in Kerala clearly show that the older population has been adversely affected by the pandemic, which is consistent with the international studies [14,24–27]. The fatality has been mostly reported from the 60-69 age category, which can be explained by using Kerala’s socio-demographic profile (Table 1 supplementary data). It is to be noted that during the entire period of the pandemic from the first death case to May 19, 2021, the median age of the death cases remains static indicating that even after the second wave of COVID-19 infection with increased mortality rates, the association between the age and fatality remains. Here it is also to be noted that the occupational structure of Kerala indicates that elderly Work Participation Rate (WPR) has considerably increased; they continue as the major contributor to the household since 1983 by working in informal, low paying occupations in poor work environments [28]. This exposure to the outside environment during the pandemic might have led them to the increased infections among the older population.

Despite these demographical factors, many older people are incapacitated due to attributable age-related diseases. This is consistent with the previous report [11] that, in the last two decades, the morbidity burden has grown faster than the rest of India wherein the Proportion of Ailing Population (PAP) in Kerala during 1995-96 was 109 against the national average of 55 which was increased to 251 in 2004, and to 308 in 2014, showing, an increase of 57 points in the overall morbidity rate against national average increase of just seven points during the 2004-2014 period. Existing evidence on the health profile suggests that Kerala has been going through a demographic transition with an unprecedented increase in the NCDs burden (Table 1). Qualitative data also validate this finding (Table VII supplementary data). This is akin to the previous study results of Kerala over a decade [8,11–13].

This disease profile (Table 1) validates the comorbidity status of the older COVID-19 death cases in Kerala (Table 2) where the most prevalent comorbidities were hypertension, diabetes, heart disease, COPD (Chronic Obstructive Pulmonary Disease) and kidney disease where many of them had multi-morbidities. Many previous studies on COVID-19 have obtained the same results [22,29–34]. To be more specific, the major comorbidity found in Kerala’s COVID-19 death cases is heart disease, followed by diabetes which are major predictors of COVID-19 fatality [35–39]. It is found that, in Kerala, the incidence of type 2 diabetes (T2DM) is 21.9% [40]. This can be attributable to risky health behaviours such as lack of exercise and an unhealthy diet which necessitates urgent implementation of healthy behaviour policy initiative. Previous studies on Middle East Respiratory Syndrome (MERS-CoV) also found comorbidity leads to MERS-CoV infection [22,41]. In West Bengal, comorbidity accounts for 90% of the COVID-19 deaths [42]. Therefore, it can be assumed that a high proportion of older people with NCDs prevalence leading to multi-morbidities can be attributed to the increased older people fatality of COVID-19 in Kerala. Nevertheless, this study discards the findings that there is no significant relationship between COVID-19 infection and diabetes [34,35] and cancer [43].

The outcome of convergence of NCDs and COVID-19 infection is serious, and it is fatal when there is multi-morbidity. Multi-morbidity gives rise to multiple interactions between one condition and the treatment recommendations for another which necessitates simultaneous multiple drug use leading to complications. The susceptibility of the older people to COVID-19 is explained by *immunosenescence* where the innate immune cells’ function is impaired when there is a decrease in the naïve T as well as B cells production, consequently leading to a situation where the innate immunity cannot fight the infection [44]. Another characteristic of ageing immunity is the CSSI (Chronic Subclinical Systemic Inflammation), which results in an elevation of inflammatory cytokines in serum due to the failure to resolve severe inflammation, which is a critical pathogenic mechanism in COVID-19, contribute to poor clinical outcome in older people [45]. This phenomenon called cytokine storm/ hypercytokinemia, associated impairment begins with the damage of the lungs’ epithelial barrier. Subsequently, this initiates a cascade of tissue damage in other vital human organs, including the heart, kidneys, brain, and blood vessels, leading to Multiple Organ Dysfunction Syndrome (MODS), which may be even more fatal [46].

Studies report that most recovered patients experienced different manifestations even 20 days after the PCR reported negative for SARS-CoV-2. The severity of such post-COVID-19 manifestations was positively correlated to the severity of the condition, which in turn was associated with the prevalence of comorbidities and age [47]. Hence, post COVID-19 clinical care and close observation of the older people are important to avoid fatalities [48]. Here is the significance of evaluation of elderly health profiles which are predictors of both COVID-19 fatality and post COVID syndrome. The situation urges to have data on the health and comorbidity profile of elderly in every country so that a clear understanding of these risk factors will help the healthcare system, particularly the clinicians, to identify and implement protocols to mitigate the fatal outcomes. Earlier, in 2013, researchers were alarmed about the threat of convergence of infectious diseases and NCDs, which will affect many in the decades to come [49]. In this context, increased investments in public healthcare facilities are necessary to ensure quality healthcare access [7], and handle post-COVID-19 syndrome situations.

Organized preventive and curative care for infectious diseases and NCDs must be ensured to the older people in the line of the ‘Vayomithram’ project of Kerala, where free health check-ups and medicines are available in proximity, particularly when Kerala is undergoing demographic transition with largest proportion of elderly in India. Geriatric care must be expanded with a pool of geriatricians in the light of possible syndemic interactions of chronic diseases and their treatments [50]. Since epidemiology has a holistic approach on wellness and maintenance, priority must be given to the complete vaccination of eldely even though 62.75 percent of the elderly are already vaccinated (first dose only) as on May 19, 2020, in Kerala. This priority in vaccination has resulted in bringing down the elderly mortality to 74.54 percent in the second wave from 76 percent in the first wave (Fig.1).

Ageing is inevitable, and often, the promotional and preventative aspects of geriatric care are neglected with a notion that it is ‘unavoidable’ and 2018;genetically determined’ and the increased disease risk is attributed to ‘normal ageing,’ neglecting the impact of healthier lifestyle to decrease healthcare expenditure [51]. To ensure geriatric care, there must be an attitudinal change and subsequent efforts from the policymakers to ensure ‘quality ageing.’ Furthermore, the public healthcare system should ensure documentation of incidents and causes of death to estimate ‘excess deaths’ occurring during outbreaks, when it becomes difficult to estimate the CFR. Weekly/ monthly death counts can be collated with trends over the years to ascertain whether it is significantly higher than the expected count. This estimation can provide information about the potential burden of mortality and fatality, associated with the infection, directly or indirectly [52].

The study finding on older people’s comorbidity level and their subsequent mortality determine the aetiology of COVID-19 and, therefore, prevention strategies can be implemented to avoid further spread and increased fatalities. Moreover, the study findings will add to the existing knowledge realm on the spectrum of comorbidities among the older people and its converged impact on the phase of epidemics spread. Future studies can be undertaken to assess the impact of vaccination after vaccinating the total older population in Kerala.

The study has its limitations. First, this study was carried out among the older people in Kerala, and therefore, the results may not be suitable in an international context. Second, getting authentic and correct primary data in calculating CFR of the elderly was a limitation. Third, for the COVID-19, as in the case of previous infectious diseases, the accurate level of transmission is often underestimated as a considerable proportion of the infected population goes undetected due to asymptomatic or mild symptomatic cases failing to test for the infection [53,54]. Fourth, there will be vulnerable segments that are either neglected or under-served and under-detected who are likely to keep themselves away from testing and treatment [55].

## CONCLUSION

It is found that older patients with chronic diseases are more susceptible to COVID-19 infection. Knowledge of these risk factors and the present health profile of the older population in Kerala necessitate a more focused approach to introduce a spectrum of interventions to protect the lives of the older people when there is a convergence of epidemics and NCDs. The study indicates the need for having timely and accurate data of a population on their demographic profile, which can easily guide the healthcare system and authorities to more efficient prevention and treatment methodologies in health care emergencies. Having evidenced the severity of COVID-19 to the older people, health commodities such as preventing vaccines and diagnostics must be made available to them at the earliest possible.

## Data Availability

The COVID-19 infection, recovery and fatality datasets generated and analyzed during the current study are available in the DHS dashboard [https://dashboard.kerala.gov.in] and the citizen-science dashboard maintained by the CODD-K consortium [https://covid19kerala.info]. Comorbidity data and death reports are available from the DHS website [https://dhs.kerala.gov.in/advisories]. Vaccination data is available from the Co-WIN website [https://www.cowin.gov.in]. The datasets on health status of older adults collected for this study are not publicly available but are available from the corresponding author upon reasonable request.

https://covid19kerala.info/

---

**Supplementary Table 1.** Socio-Demographic Variables.

| **Variable** | **Category** | **Frequency** | **Percent** |
| --- | --- | --- | --- |
| Gender | Female | 181 | 44.7 |
|  | Male | 224 | 55.3 |
| Age | 60-69 | 203 | 50.1 |
|  | 70-79 | 128 | 31.6 |
|  | 80-89 | 60 | 14.8 |
|  | Above 90 | 9 | 2.2 |
| Marital Status | Married | 314 | 77.5 |
|  | Not Married | 11 | 2.7 |
|  | Partner Died | 75 | 18.5 |
|  | Divorced | 3 | 0.7 |
| Education | No formal Education | 74 | 18.3 |
|  | School | 267 | 65.9 |
|  | College/University | 53 | 13.1 |
|  | No response | 4 | 1 |
| Monthly income | No income | 125 | 25.9 |
|  | Less than INR 10,000 | 228 | 56.3 |
|  | INR 10,000-20,000 | 37 | 9.1 |
|  | INR 21,000-30,000 | 20 | 4.9 |
|  | Above INR 30,000 | 5 | 1.2 |
| Source of Income | Job | 18 | 4.4 |
|  | Pension | 211 | 52.1 |
|  | Income from assets | 29 | 7.2 |
|  | Business | 35 | 8.6 |
|  | Income from agriculture | 49 | 12.1 |
|  | Income from other sources | 15 | 3.7 |

**Supplementary Table 2.** Disease and health behaviour of elderly - Qualitative data.

| Attribute | Responses (District-participant ID, Gender, Age) |
| --- | --- |
| Disease | ..I have Diabetes for the past 35 years, severe joint pain, back pain... (Palakkad, F, 70-75)<br>...there are number of diseases in this age...headache, back pain, blood pressure, disk problem, cholesterol (Pathanamthitta, F, 60-65)<br>I am taking medicines for blood pressure regularly.... (Pathanamthitta, M, 60-65)<br>....I had a block in my heart, for which cardiac surgery was done. Check-up is done every three months. Then I had prostate cancer... follow up visits once in six months. I am a diabetic patient too... (Thrissur, M, 65-70)<br>...angioplasty was done ....taking medicines for blood pressure twice a day.” (Kozhikode, F, 70-75)<br>“I am a cardiac patient, using a pacemaker.... (Palakkad, F, 70-75)<br>“Taking medicines for thyroid, sugar, BP daily... (Thiruvananthapuram, F, 65-70)<br>“My current health problems are Asthma, Cholesterol, Sugar, blood pressure, thyroid, Cardiac diseases..... (Kollam, F, 70-75) |
|  | I have cancer, BP, Heart problem, doing chemotherapy<br>(Malappuram, M, 70-75) |

